# Effectiveness of vaccines in preventing hospitalization due to COVID-19: A multicenter hospital-based case-control study, Germany, June 2021 to January 2022

**DOI:** 10.1101/2022.06.28.22276303

**Authors:** Anna Stoliaroff-Pepin, Caroline Peine, Tim Herath, Johannes Lachmann, Delphine Perriat, Achim Dörre, Andreas Nitsche, Janine Michel, Marica Grossegesse, Natalie Hofmann, Thomas Rinner, Claudia Kohl, Annika Brinkmann, Tanja Meyer, Brigitte G. Dorner, Daniel Stern, Fridolin Treindl, Sascha Hein, Laura Werel, Eberhard Hildt, Sven Gläser, Helmut Schühlen, Caroline Isner, Alexander Peric, Ammar Ghouzi, Annette Reichardt, Matthias Janneck, Guntram Lock, Lars Schaade, Ole Wichmann, Thomas Harder

**Author notes:** equal contribution (Anna Stoliaroff-Pepin, Caroline Peine).

## Abstract

We included 852 patients in a prospectively recruiting multicenter matched case-control study in Germany to assess vaccine effectiveness (VE) in preventing COVID-19-associated hospitalization (Delta-variant dominance). Two-dose VE was 89% (95%CI 84-93%) overall, 79% in patients with >2 comorbidities and 77% in adults aged 60-75 years. A third dose increased VE to >93% in all patient-subgroups.

## Background

In spring 2021, a new variant of concern (VOC), the highly contagious SARS-CoV-2 Delta variant, was identified and gained global dominance by summer 2021. As of August 2021, the weekly incidence in Germany steadily increased, reaching more than 300,000 newly infected persons per week by November 2021. A combination of non-pharmaceutical interventions and a booster vaccination campaign (three vaccine doses) led to decreasing case numbers until the Omicron variant of SARS-CoV-2 took over in 2022 [1].

High efficacy of COVID-19 vaccines has been demonstrated in randomized, placebo-controlled clinical trials (73% for vector vaccines and 85% for mRNA vaccines in meta-analyses) [2], yet real world data may differ depending on population characteristics and the dominant viral strain during the observation period. In addition, heterologous vaccination schemes i.e. using a vector vaccine for the first and an mRNA vaccine for the second injection, or vice versa, commonly occurred. For booster vaccination, only mRNA vaccines (Comirnaty®, Spikevax®) were recommended in Germany. Furthermore, there is limited data on the duration of protection under the Delta-variant [3-5].

Early in 2021, we set up a study in 13 hospitals across Germany to analyze the effectiveness of the COVID-19 vaccines in Germany in preventing COVID-19 associated hospitalization in the adult population. In addition, we aimed to perform subgroup analyses for vaccine effectiveness (VE) by age, sex, severity of disease, underlying comorbidities and time since vaccination. Here we present VE results assessed during the Delta-variant wave.

## Methods

COViK is a prospectively recruiting matched case-control study in Germany that is led by Germany’s Public Health Institute (Robert Koch Institute, RKI). Study sites are 13 hospitals in Berlin, Hamburg, Wuppertal, Duesseldorf, Erfurt and Chemnitz. The study was commenced on 1 June 2021 and is expected to end in June 2023.

We here present an interim analysis based on the data of patients recruited from 1 June 2021 until 31 January 2022. A case was defined as an adult aged 18 to 90 years who had a positive SARS-CoV-2 PCR test and was hospitalized in one of the 13 study hospitals. To be included, cases had to be either hospitalized due to a severe COVID-19 infection or COVID-19 complications, or with a severe nosocomial COVID-19 infection (see supplementary material). Controls were hospitalized patients who were tested negative for SARS-CoV-2 by PCR and were recruited at surgical, orthopaedical, urological and gynecological wards, preferably with acute diseases (e.g. fracture, tendon rupture). Two controls per case were recruited from the same hospital that the respective case was admitted to, or, if not available, from a hospital in the same city (1:2 matching). Controls were matched to cases based on their hospital admission date (+/-14 days), age (+/-10 years) and sex. All study nurses were trained by the COVIK study centre team regularly. Repeated on-site visits and quality controls were performed to ensure adherence to the study protocol.

Cases and controls with a previous SARS-CoV-2 infection were excluded from the analysis to avoid misclassification (e.g. risk of infection, indication for vaccination; Supplementary Material Table 1).

A current SARS-CoV-2 infection was identified through a positive result of a SARS-CoV-2-real-time PCR performed on a naso-oropharyngeal swab [6]. The virus variant was determined by sequencing using the AmpliCoV protocol [7], or by PCR typing assays if RNA load was too low for sequencing. In addition, antibodies against SARS-CoV-2 and other coronaviruses were determined and virus-neutralization tests were performed for future immunological analyses.

Data on basic socio-demographic factors (e.g. age, sex, level of graduation), COVID-19 disease (e.g. previous infection, date of symptom onset), COVID-19-vaccination (e.g. vaccination status, vaccine type and number of vaccine doses administered), risk factors for COVID-19 infection and for severe course of disease (e.g. comorbidities) were collected in individual interviews conducted by research nurses. Clinical and laboratory data were extracted from medical records (e.g. sequencing results, admission to an intensive care unit (ICU)). All participants (or their legal guardians) provided a written informed consent to participate in the study.

For this first pre-specified interim analysis, we computed the 2-dose and 3-dose VE regardless of the individual matching for the following subgroups: males and females, aged 18-59, 60-75, and 76-90 years, with <3 or ≥ 3 pre-existing comorbidities, last vaccine dose administered in the past 3 months or 3-6 months ago, admitted to intensive care or not (Figure 1, Supplementary Table 3). Patients vaccinated only once were excluded from the analysis of vaccine effectiveness. The analysis was restricted to cases infected with the Alpha or Delta variant.

**Figure 1:**
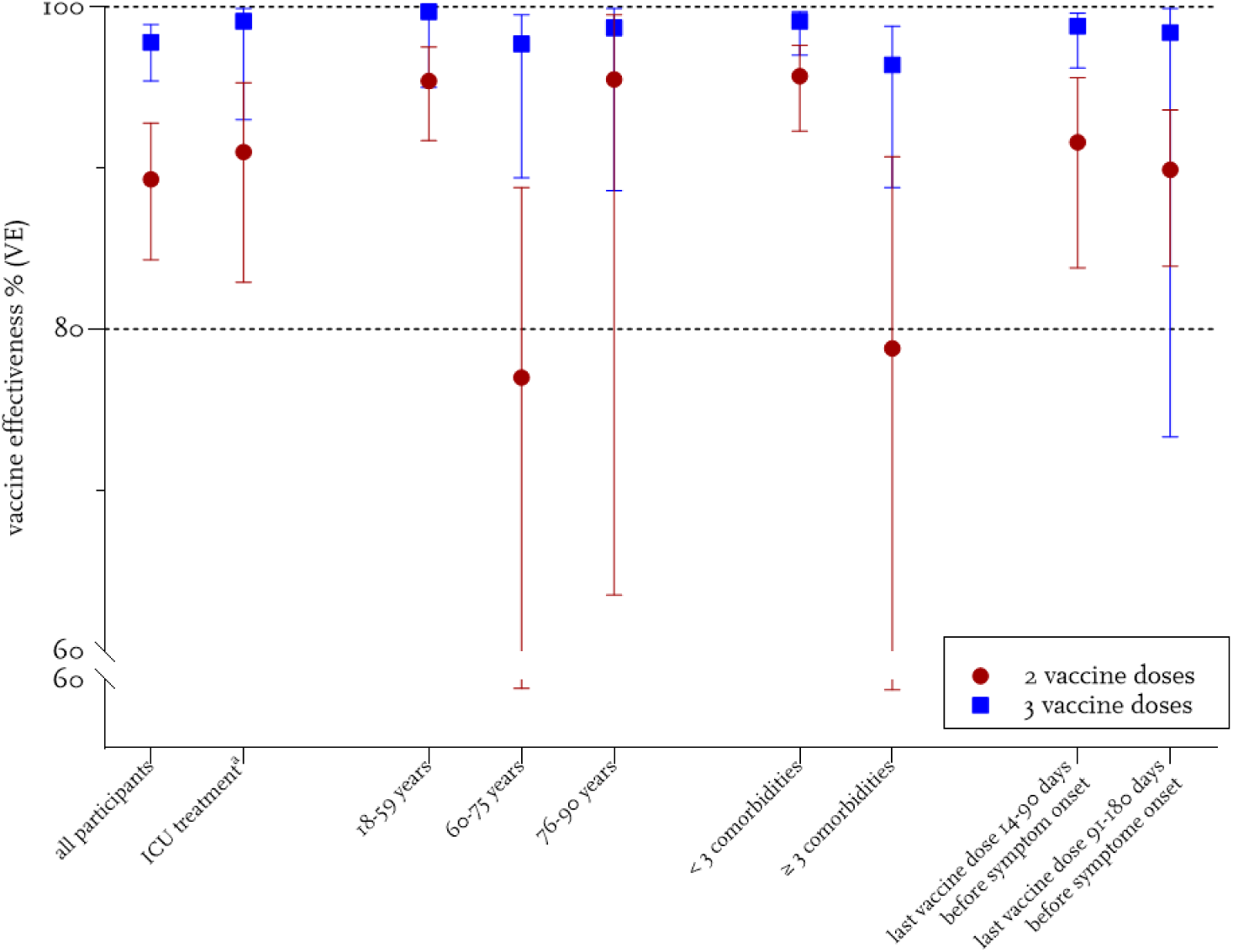
Vaccine effectiveness for two and three vaccination doses, endpoint severe COVID-19 (hospitalization) ^a^ICU treated cases versus controls with no ICU treatment

Based on logistic regression modeling, we determined VE, adjusted for age, education and pre-existing comorbidities.

Comparison of the case and control groups in Supplementary Table 2, was performed with appropriate significance tests (t-tests for continuous variables and chi-squared tests or Fisher’s exact test for categorical variables; see Supplementary Material).

Data were analyzed using the statistical software R, version 4.1.2.

The study was approved by the Ethics Committee of the Charité Universitätsmedizin, Berlin (EA1/063/21) and was registered at “Deutsches Register Klinischer Studien” (DRKS00025004).

## Results

During the study period, 852 participants were recruited, including 244 cases and 608 matched controls.

Median age of cases was 57 years (interquartile range 45-70 years), median age of controls was 59 years (interquartile range 48-72 years), 45 % of cases and 43% of the controls were female. Cases had a lower education status than controls (42% of cases vs 28% of controls with nine or less school years, (p< 0.05)). The majority of the cases was infected with the Delta variant (79%), and 5% of them were infected with the Alpha variant (missing information 16%). Nearly a quarter of cases (n=56, 23%) was admitted to the ICU and 13 patients (5,3%) deceased (Supplementary Table 2).

Cases were significantly less often vaccinated against COVID-19 than controls: More than half of the cases (58%, n=142/244) were not vaccinated at all, compared to 11% of the controls. Nearly a third (30%, 73/244) of the cases and more than half of the controls (53%, 323/608) had received two vaccine doses (first vaccination series) and 9/244 cases (4%) and 192/316 controls (32%) had received a third dose (booster dose). Additionally, three controls had received a fourth vaccine dose.

The most frequently administered vaccine among study participants was Comirnaty® (Biontech/Pfizer), 1119 doses, followed by Vaxzevria® (Astra-Zeneca, 151 doses) and Spikevax® (Moderna, 141 doses), while Jcovden® (Johnson& Johnson/Janssen) was administered 28 times.

After adjustment for age, education and pre-existing comorbidities, overall VE (all groups) was 93.5% (CI 89.1 – 96.2%) after two vaccine doses and 99.4% (CI 98.1-99.9%) for three doses.

VE after two vaccine doses was significantly lower for adults with three or more pre-existing comorbidities as compared to adults with less than three comorbidities (95.7% vs 78.7%), while VE after three vaccine doses was similar for both groups (Figure 1, Supplementary Table 3).

The VE was lower among adults aged 60-75 years but this reduction was again compensated when a third dose was administered. When the individual matching of the pairs was considered, similar results were obtained (Supplementary Table 4).

## Discussion

To our knowledge, this is the first study in Germany that assessed VE of COVID-19 vaccines. Our analysis suggests that the COVID-19-vaccines licensed in the European Union were highly effective in preventing hospitalization due to COVID-19.

The vaccine effectiveness was 93.5% after two vaccine doses and 99.4% after three doses. Recently, studies from Canada, the UK and others have reported comparably high VE after two doses against hospitalization [4] with a slow decrease in protection against hospitalization over time. Importantly, waning of protection especially affected clinically vulnerable groups [8]. We found that vaccination protected from severe disease for at least six months and the moderately reduced two-dose protection after three to six months (90%) raised again to 98% after a booster shot, emphasizing the necessity of the third vaccine dose.

Patients between 60 and 75 years of age had a significantly reduced two-dose effectiveness, probably due to a weaker immune response upon vaccination compared with younger people. This group benefitted in particular from a third dose. Almost a quarter of our study participants was admitted to an ICU. However, this number may underestimate the ratio of very severe cases as some patients, e.g. elderly people, may refuse ICU treatment.

### A main advantage of our study design stems from the ability to collect detailed high-quality information in a prospective manner

Every COVID-19 diagnosis was confirmed by clinical records and -if necessary-by direct consultation of the attending physician. Only patients requiring hospitalization due to COVID-19 were included. Many post-deployment studies rely on clinical data registries, resulting in fast reporting which is valuable to guide policy decisions during a pandemic. However, not all patients are included in such registries and their main diagnosis may not be COVID-19. Unlike many other studies, we explicitly examined the pre-existing immunity by serology early in the course of infection and excluded all participants with pre-existing antibodies or with a previous laboratory-confirmed SARS-CoV-2 infection. The detailed verification leads to a better data quality: With relying on patient’s information only we would have excluded six patients due to previous SARS-CoV-2 infection, the antibody test revealed 24 further cases. Participants with a history of SARS-CoV-2 infection were excluded from the analysis to avoid a biased analysis as previous infections potentially prevent subsequent infections and vaccination was not recommended at least for three months after infection after SARS-CoV-2 infection in Germany. Furthermore, we determined the virus variant for each patient.

A limitation of our study is the low number of participants. We could not gain precise results for matched pairs and triplet analysis for some subgroups for this reason.

This first interim analysis provides encouraging results and warrants follow-up analyses to assess the evolving COVID-19 vaccine effectiveness, in a changing epidemiologic landscape in terms of circulating variants, available vaccines, and increasing population-wide immunity. In the future course of the study, we plan to analyze the Omicron wave, combinations of natural infection and vaccination, longer time intervals since vaccination, long-term COVID-19 symptoms in vaccinated vs unvaccinated individuals (long COVID), and the immune response after COVID-19 breakthrough infections.

## Conclusion

The COVID-19 vaccines were highly protective against hospitalization in real-world settings in Germany during the Delta-variant predominance. Reduced vaccine effectiveness observed in subgroups after two doses was compensated after three doses. This finding would support efforts to maximize vaccine uptake to three doses among vulnerable populations.

## Data Availability

All data produced in the present study are available upon reasonable request to the authors

## Acknowlegments

The authors thank all study nurses for the valuable contribution, namely Sawsanh Al-Ogaidi, Nancy Beetz, Belgin Esen, Rola Khalife, Marie-Kristin Kusnierz, Katja Lange, Luise Mauer, Antje Micheel, Marlies Schmidt, Yvonne Weis, Franziska Weiser, Aysete Yencilek. We thank Wiebke Hellenbrand for her important contribution in designing and setting up the study as well as Anna Meier, Swetlana Muminow, Richard Schensar, Ellen Busch, Hanna Buck, and Moritz Gehring for their support in organizing the project and Vincent Stoliaroff-Pépin for support with R.

## Supplementary Material

**Supplementary Table 1.**
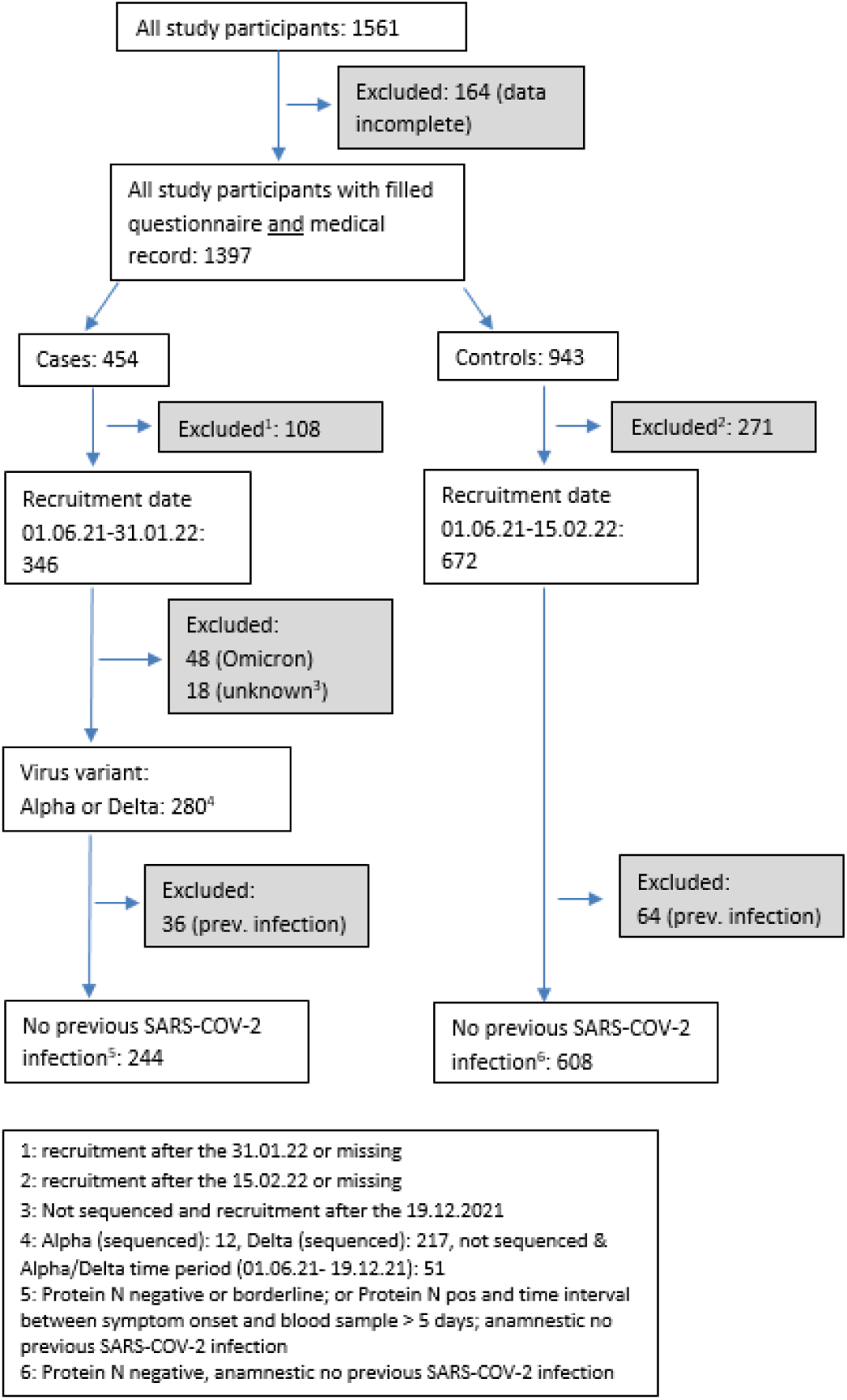
Inclusion /exclusion criteria.

**Supplementary Table 2.** Patient characteristics.

|  | case | control | p-value |
| --- | --- | --- | --- |
| **n** | 244 | 608 |  |
| **Age (years) mean (SD)** | 57.09 (16.6) | 58.96 (15.4) | 0.119 |
| **Age group n (%)** |  |  | 0.039 |
| 18-59 years | 139 (57.0) | 298 (49.0) |  |
| 60-75 years | 62 (25.4) | 208 (34.2) |  |
| 76-90 years | 43 (17.6) | 102 (16.8) |  |
| **Sex n (%)** |  |  | 0.731 |
| male | 135 (55.3) | 346 (56.9) |  |
| female | 109 (44.7) | 262 (43.1) |  |
| **Highest school educational level n (%)** |  |  | <0.001 |
| no graduation | 53 (21.7) | 26 (4.3) |  |
| 9 school years | 49 (20.1) | 142 (23.4) |  |
| 10 school years (secondary school certificate) | 77 (31.6) | 228 (37.5) |  |
| 12 or 13 school years (high school graduation) | 65 (26.6) | 212 (34.9) |  |
| **BMI (kg/m<sup>2</sup>) mean (SD))** | 28.65 (7.04) | 27.19 (12.50) | 0.086 |
| **BMI group n (%)** |  |  | <0.001 |
| underweight/normal weight (BMI ≤ 25 kg/m<sup>2</sup>) | 74 (30.3) | 266 (43.8) |  |
| overweight (BMI > 25-30 kg/m<sup>2</sup>) | 82 (33.6) | 198 (32.6) |  |
| obesity (BMI > 30 kg/m<sup>2</sup>) | 88 (36.1) | 144 (23.7) |  |
| **Admission to intensive care unit (ICU) n (%)** |  |  |  |
| yes | 56 (23.0) |  |  |
| no | 188 (77.0) |  |  |
| **Death n (%)** |  |  |  |
| yes | 13 (5.3) |  |  |
| no | 227 (93.0) |  |  |
| NA | 4 (1.6) |  |  |
| **Pre-existing comorbidities (general) n (%)** |  |  | <0.001 |
| ≤ 2 | 121 (49.6) | 411 (67.6) |  |
| > 2 | 98 (40.2) | 112 (18.4) |  |
| NA | 25 (10.2) | 85 (14.0) |  |
| **Pre-existing comorbidities (immune system) n (%)** |  |  | 0.161 |
| non | 211 (86.5) | 500 (82.2) |  |
| ≥ 1 | 33 (13.5) | 108 (17.8) |  |
| **Pre-existing comorbidities (high risk for severe COVID-19)<sup>a</sup> (%)** |  |  | 0.143 |
| ≤ 2 | 231 (94.7) | 590 (97.0) |  |
| >2 | 13 (5.3) | 18 (3.0) |  |
| **Vaccination (≥ 2 doses) n (%)** |  |  |  |
| yes | 82 (33.6) | 518 (85.2) | <0.001 |
| no | 162 (66.4) | 90 (14.8) |  |
| **Number of vaccine doses n (%)** |  |  |  |
| 0 | 142 (58.2) | 67 (11.0) | <0.001 |
| 1 | 20 (8.2) | 23 (3.8) |  |
| 2 | 73 (29.9) | 323 (53.1) |  |
| 3 | 9 (3.7) | 192 (31.6) |  |
| 4 | 0 (0.0) | 3 (0.5) |  |
| **Vaccine type n (%)** |  |  |  |
| mRNA | 12 (4.9) | 12 (2.0) |  |
| vector | 8 (3.3) | 11 (1.8) |  |
| mRNA/mRNA | 62 (25.4) | 255 (41.9) |  |
| vector/vector | 7 (2.9) | 27 (4.4) |  |
| crossover<sup>b</sup> | 3 (1.2) | 36 (5.9) |  |
| mRNA/mRNA/mRNA | 7 (2.9) | 157 (25.8) |  |
| mRNA/mRNA/vector | 0 (0.0) | 1 (0.2) |  |
| vector/vector/vector | 0 (0.0) | 0 (0.0) |  |
| vector/vector/mRNA | 1 (0.4) | 13 (2.1) |  |
| crossover<sup>b</sup>/vector | 0 (0.0) | 1 (0.2) |  |
| crossover<sup>b</sup>/mRNA | 0 (0.0) | 18 (3.0) |  |
| NA | 2 (0.8) | 6 (1.0) |  |

<sup>a</sup> High-risk comorbidities for severe course of COVID-19: diabetes type 2, Organ transplant, BMI> 30, COPD, renal insufficiency, heart failure[12]
<sup>b</sup> first dose mRNA, second dose vector vaccine or vice versa

**Supplementary Table 3.** COVID-19 vaccine effectiveness (VE) against hospitalization after two or three vaccine doses, interim analysis of the COVIK study, June 2021-January 2022, Germany.

|  | Calculation of 2-dose VE (vs 0 doses):<br>215 cases and 390 controls |  |  | Calculation of 3-dose VE (vs 0 doses):<br>151 cases and 259 controls |  |  | 3 vs. 2 vaccine doses |
| --- | --- | --- | --- | --- | --- | --- | --- |
|  | Vaccinated<br>cases<br>n (%) | Vaccinated<br>controls<br>n (%) | VE (%)<br>(95% CI) | Vaccinated<br>cases<br>n (%) | Vaccinated<br>controls<br>n (%) | VE (%)<br>(95% CI) | VE (%) (95%CI) |
| **All patients** | 73/215 (34.0) | 323/390 (82.2) | 89.3 (84.3-92.8) | 9/151 (6.0) | 192/259 (74.1) | 97.8 (95.4-98.9) | 79.3 (57.6-89.9) |
| **Sex** |  |  |  |  |  |  |  |
| Male | 41/115 (35.7) | 168/211 (79.6) | 85.8 (76.4-91.5) | 6/80 (7.5) | 115/158 (72.8) | 97.0 (92.5-98.8) | 78.6 (48.0-91.2) |
| Female | 32/100 (32.0) | 155/179 (86.6) | 92.7 (86.7-96.0) | 3/71 (4.2) | 77/101 (76.2) | 98.6 (95.2-99.6) | 81.1 (36.4-94.4) |
| **Age (years)** |  |  |  |  |  |  |  |
| 18-59 | 17/124 (13.7) | 164/211 (77.7) | 95.4 (91.7-97.5) | 0/107 | 73/120 (60.8) | 99.7 (95.0-100) | NA |
| 60-75 | 32/56 (57.1) | 110/129 (85.3) | 77.0 (52.7-88.8) | 2/26 (7.7) | 69/88 (78.4) | 97.7 (89.4-99.5) | 90.0 (57.1-97.7) |
| 76-90 | 24/35 (68.6) | 49/50 (98.0) | 95.5 (63.5-99.5) | 7/18 (38.9) | 50/51 (98.0) | 98.7 (88.6-99.9) | 71.4 (27.6-88.7) |
| **Pre-existing**<br>**comorbidities<sup>a</sup>** |  |  |  |  |  |  |  |
| < 3 | 20/107 (18.7) | 210/249 (84.3) | 95.7 (92.3-97.6) | 3/90 (3.3) | 150/189 (79.4) | 99.1 (97.0-99.7) | 79.0 (28.1-93.9) |
| ≥ 3 | 49/86 (57.0) | 56/65 (86.2) | 78.7 (51.5-90.7) | 6/43 (14.0) | 40/49 (81.6) | 96.4 (88.8-98.8) | 82.9 (56.1-93.3) |
| **Time between**<br>**last vaccine dose and**<br>**symptom onset<sup>b</sup> (days)** |  |  |  |  |  |  |  |
| 14-90 | 13/155 (8.4) | 73/140 (52.1) | 91.6 (83.8-95.6) | 3/145 (2.1) | 122/189 (64.6) | 98.8 (96.2-99.6) | 86.2 (49.9-96.2) |
| 91-180 | 37/179 (20.6) | 172/239 (72.0) | 89.9 (83.9-93.6) | 0/142 | 15/82 (10.5) | 98.4 (73.3-99.9) | 84.5 (165.2-99.1) |
| **ICU admission<sup>c</sup>** |  |  |  |  |  |  |  |
| Yes | 16/53 (30.2) | 323/390 (82.8) | 91.0 (82.9-95.3) | 1/38 (2.6) | 192/260 (73.8) | 99.1 (93.0-99.9) | 89.5 (20.1-98.6) |
| No | 57/162 (35.2) | 323/390 (82.8) | 88.7 (82.9-92.6) | 8/113 (7.1) | 192/260 (73.8) | 97.3 (94.3-98.8) | 76.4 (49.5-89.0) |

Hospitalization VE: COVID-19 Vaccine Effectiveness against hospitalization; 95% CI: 95% Confidence Interval; ICU: Intensive Care Unit; NA: Not Applicable; <sup>a</sup> Examples of pre-existing comorbidities: Heart failure, renal insufficiency, diabetes mellitus, COPD, stroke; <sup>b</sup> for controls: time between the last vaccine dose and hospital admission; <sup>c</sup> cases with ICU treatment versus controls without ICU admission treatment

### 4. Supplementary Methods Sequencing/ PCR typing

Samples were taken nasopharyngeally and oropharyngeally, stored in a refrigerator at 2-8°C, and sent by post with attached cold packs or courier twice a week. All laboratory analyses were performed in the Centre for Biological Threats and Special Pathogens at the Robert Koch-Institute in Berlin. If whole genome sequencing was not possible, the variant was identified by PCR typing. If the virus detection by PCR was not successful due to low RNA load, the study nurse was asked to send a sputum sample to the RKI laboratory. If the sputum could not be sequenced, the sequencing result of the hospital was used if available.

#### Inclusion/exclusion criteria and matching of controls

Cases had to be tested PCR-positive to be included. Only positive tests that were performed directly before, at, or directly after the admission and were temporally related to the current infection were assessed. Only patients with severe COVID-19 were included in the study as cases. A clinical case definition was not applied as the symptoms of COVID-19 including all complications can vary from case to case and, over time, and also from variant to variant. All patients whose COVID-19 symptoms were too severe to be treated as outpatients and who were hospitalized for COVID-19 were eligible. In patients with pre-existing comorbidities it was sometimes difficult to decide whether hospitalization occurred because of COVID-19 or whether it was mainly related to comorbidities. If the degree of severity of COVID-19 was ambiguous in patients with pre-existing comorbidities, the physician in charge was consulted, to decide whether COVID-19 (alone or by deterioration of pre-existing comorbidities) of the potential case was so severe that the patient had to be hospitalized for this reason.

Controls with a previous SARS-CoV-2 infection were excluded independently from the date of infection. A previous SARS-CoV-2 infection was defined as a positive result of an in-house established multiplex assay performed on a serum sample for the detection of N-protein antibodies. N-protein positive cases with blood sampling within five days after symptom onset were excluded too. Additional all cases and controls with a reported history of a previous SARS-CoV-2 infection were excluded.

Cases with Omicron variant infections were excluded from the current analysis. Cases with missing sequencing results were considered to belong to the Alpha/Delta group until December 19, 2021, as Delta was the dominant variant (97 % Delta variant cases in Germany in calendar week 50, https://www.rki.de/DE/Content/InfAZ/N/Neuartiges_Coronavirus/Situationsberichte/Wochenbericht/Wochenbericht_2021-12-23.pdf?blob=publicationFile). Cases and controls with contraindication for a SARS-CoV-2 vaccination, e.g. anaphylactic reaction, were not included.Control patients were carefully selected to represent the general population. Study nurses were asked to prioritize the inclusion of non-elective patients. In addition, if female cases were pregnant, pregnant controls were matched.

#### Changes due to dynamic pandemic situation, adjustment of study design

The dynamic situation in the pandemic made adjustments of the study design at some points necessary. Due to a lack of vaccine, prioritized groups, e.g. health care workers, elderly, and high-risk patients, were vaccinated in the first line in spring 2021 in Germany. We therefore excluded non-prioritized potential participants at the beginning of the study, because their chance of being vaccinated was very low at that time. Furthermore, we excluded potential participants with a history of SARS-CoV-2 in the analysis of the delta wave, but we decided recently to include these participants in our next analysis (omicron wave), as the SARS-CoV-2 prevalence became high meanwhile and the exclusion of recovered controls may lead to overestimation of VE as recovered patients have a lower likelihood to be vaccinated. However, studies comparing controls with or without previous infection reported similar results between the different types of controls [9].

With the increase of vaccine coverage in the population, the characteristics of unvaccinated people change. At a high vaccination coverage, they form a special group [10, 11]. However, as this applies to cases as well as controls, we do not expect a significant bias in this regard. The unvaccinated people in population with high vaccination coverage may also have a higher infection risk as they might not adhere to non-pharmaceutical interventions, and they may more often refuse to take part in a study, which became noticeable in spring 2022. We took countermeasures in distributing incentives for our participants and offered training courses for our study nurses in terms of participant recruitment.

#### Statistical analysis

Comparison of the case and control groups was performed with appropriate significance tests (t-tests for age and BMI and chi-squared tests or Fisher’s exact test for age group, sex, educational level, comorbidities, vaccination status and number of vaccine doses). Although our study design is a matched case-control study an unmatched analysis can be performed in the given situation [13]. As the matched subgroup-analysis was not applicable for all subgroups due to insufficient number of patients in the strata, we primarily performed an unmatched analysis. The odds ratio (OR) regarding vaccination and severe Covid-19 was calculated with the formula

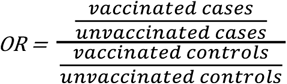

Based on this, the VE regarding severe Covid-19 was calculated with the formula

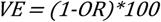

For estimating the VE to prevent ICU admission, for non-ICU patients and with regard to protection for time intervals after the last vaccination the calculation was performed across subgroups.

Furthermore, pairwise matched analysis was performed, to confirm the robustness of results (Supplementary Table 4). Pairwise matched analysis was calculated by the Mantel-Haenszel method for matched data on the basis of discordant pairs (see Miettinen (1970)). In some of the subgroup analyses, e.g. boostered ICU patients we applied the Woolf-Haldane correction as the combination vaccinated case/unvaccinated control(s) was not present in every subset of data.

**Supplementary Table 3.** Results of pairwise matched analysis vs unmatched analysis.

|  | Unmatched analysis | Unmatched analysis | Matched analysis | Matched analysis |
| --- | --- | --- | --- | --- |
|  | VE after 2 doses<br>(95% CI) | VE after 3 doses<br>(booster)<br>(95% CI) | VE after 2 doses<br>(95% CI) | VE after 3 doses<br>(booster)<br>(95% CI) |
| **All patients** | 89.3% (84.3-92.8%) | 97.8% (95.4-98.9%) | 93.4% (92.8-94.0%) | 98.1% (97.4-98.7%) |
| **Male** | 85.8 (76.4-91.5%) | 97.0% (92.5-98.8%) | 89.1% (87.5-90.5%) | 97.7% (96.6-98.4%) |
| **Female** | 92.7% (86.7-96.0%) | 98.6% (95.2-99.6%) | 95.3% (95.5-96.1%) | 96.7% (94.1-98.1%) |
| **18-59 years** | 95.4% (91.7-97.5%) | 99.7% (95.0-100%) | 98.4% (98.1-98.6%) | 95.4% (95.9-98.3%) |
| **60-75 years** | 77.0% (52.7-88.8%) | 97.7% (89.4-99.5%) | 78.6% (64.2-87.2%) | NA |
| **76-90 years** | 95.5% (63.5-99.5%) | 98.7% (88.6-99.9%) | NA | 90.0% (24.1-98.7%) |
| **< 3 comorbidities** | 95.7% (92.3-97.6%) | 99.1% (97.0-99.7%) | 92.4% (91.4-93.8%) | NA |
| **≥ 3 comorbidities** | 78.7% (51.5-90.7%) | 96.4% (88.8-98.8%) | NA | NA |

Logistic regression was performed for 2 times vaccinated and 3 times vaccinated patients. Adjustment variables were age, school education and pre-existing comorbidities. Variable selection was performed on the basis of predictive accuracy, the AIC and context-related information.

